# Mass molecular testing for COVID19 using NGS-based technology and a highly scalable workflow

**DOI:** 10.1101/2020.08.10.20172106

**Authors:** Fernanda de Mello Malta, Deyvid Amgarten, Felipe Camilo Val, Rubia Anita Ferraz Santana, Murilo Castro Cervato, Bruna Mascaro Cordeiro de Azevedo, Marcela de Souza Basqueira, Camila Oliveira dos Santos Alves, Maria Soares Nobrega, João Renato Rebello Pinho

**Author notes:** Authors contributed equally to this work (Fernanda de Mello Malta, Deyvid Amgarten). Address for correspondence: João Renato Rebello Pinho, Laboratorio de Técnicas Especiais, Hospital Israelita Albert Einstein, Av. Albert Einstein, 627/701 05651-901 São Paulo-SP Brazil.

## Abstract

Since first reported case of the new coronavirus infection in Wuhan, China, researchers and governments have witnessed an unseen rise in the number of cases. Thanks to the rapid work of Chinese scientists, the pathogen now called SARS-CoV-2 has been identified and its whole genome has been deposited in public databases by early January 2020. The availability of the genome has allowed researchers to develop Reverse Transcription - Polymerase Chain Reaction (RT-PCR) assays, which are now the gold-standard for molecular diagnosis of the respiratory syndrome COVID19. Because of the rising number of cases and rapid spreading, the world has been facing a shortage of RT-PCR supplies, especially the ones involved in RNA extraction. This has been a major bottleneck to increase testing capacity in many countries that do not significantly manufacture these supplies (Brazil included). Additionally, RT-PCR scalability is highly dependent on equipment that usually perform testing of 96 samples at a time. In this work, we describe a cost-effective molecular NGS-based test for diagnosis of COVID19, which uses a single-step RNA extraction and presents high scalability and accuracy when compared to the gold-standard RT-PCR. A single run of the NGS-based test using the Illumina NextSeq 550 mid-end sequencing equipment is able to multiplex 1,536 patient’s samples, providing individual semi-qualitative results (detected, not detected). Detected results are provided with fragments per million (FPM) values, which was demonstrated to correlate with RT-PCR Cycle Threshold (CT) values. Besides, usage of the high-end Illumina Novaseq platform may yield diagnostic for up to 6,144 samples in a single run. Performance results when compared with RT-PCR show general accuracy of 96% (or 98% when only samples with CT values for gene N lower than 30 are considered). We have also developed an online platform, called VarsVID®, a Varstation® feature, to help test executors to easily scale testing numbers. Sample registering, wet-lab worksheets, sample sheet for sequencing and results’ display are all features provided by VarsVID® on Varstation®. Altogether, these results will contribute to control COVID19 pandemics.

## 1. Introduction

In December 2019, it was reported in Wuhan City, Hubei province in China, cases of pneumonia of unknown etiology. In 7th January 2020, the causative agent was identified from throat swab samples in a study conducted by the Chinese Center for Disease Control and Prevention (CCDC) (1). The new coronavirus was provisionally named 2019 novel coronavirus (2019-nCOV) (2) (3). Coronaviruses, from the family *Coronaviridae*, are enveloped single-strand RNA viruses with genomes ranging from 26 to 32 kilobases in size. Most members of the coronaviruses family have similar genome organization and expression, usually composed by ten or more nonstructural proteins (nsp proteins) and by the structural proteins spike (S), envelope (E), membrane (M), and nucleocapsid (N) (3). Through Next-Generation Sequencing technology (NGS), the virus complete genome was described and phylogenetically classified as belonging to the genus betacoronavirus. Thereafter, 2019-nC0V was named SARS-CoV-2 (1, 4).

On March 2020, the WHO (World Health Organization) declared COVID-19 as pandemic (5) and have issued guidelines about clinical and epidemiological findings, stating that extensive laboratory tests should be performed (6). Since then, there has been a robust scientific response and a global search for the best diagnostic tool to identify patients infected by SARS-CoV-2. Several laboratory methods have been used to detect the etiological agent of this respiratory syndrome, as for instance enzyme-linked immunosorbent assay and nucleic acid hybridization. Moreover, early publication of SARS-CoV-2 genome has allowed development of molecular tests based on reverse transcription polymerase chain reaction, widely known as RT-PCR. Months into the pandemics now, RT-PCR has been the gold standard technique for molecular identification of the virus and for diagnosis of COVID19.

NGS technology have revolutionized the genomics field and have been providing a novel and effective way to screen samples and detect pathogens. In association with bioinformatics tools, this technology is changing the way as research and diagnostic centers can respond to infectious disease outbreaks. This progress is paving the way for new approaches and for improving our knowledge about the disease origin, occurrence and transmission, through a large-scale genomic approach. Over 16,000 SARS-CoV-2 genome sequences have been deposited in public repositories until May 2020, and genome sequencing is helping researchers to understand SARS-CoV-2 haplotypes distribution, mutations and phylogeny (7-9).

In a pandemic scenario, it is essential to carry out population mass testing to prevent rapid spread of the infection and to minimize mortality rates. In response to this need, we have developed an NGS-based diagnostic test for the new coronavirus, which presents 96% of accuracy in paired analyses with the RT-PCR gold standard. Our NGS-based test is highly scalable, having the ability to perform up to 1,536 samples simultaneously on a mid-end NextSeq 550 Illumina sequencing equipment. This number is 16 times more than what is done today by the gold standard RT-PCR technique and it may be even more expanded considering the use of a high-end Novaseq 6000 Illumina equipment (3072 to 6144 samples in a single run). This new technology expands the global diagnostic capacity, enabling rapid initiation of treatment and isolation of the infected, altogether contributing to control COVID19 pandemics.

## 2. Methods

### 2.1. General workflow

We have developed a complete workflow to execute the NGS-based testing, including wet-lab and bioinformatics analyses, as shown in Figure 1. This workflow is highly scalable in order to avoid bottlenecks and to increase testing capacity. Wet-lab steps may be automated by pipetting robots, and bioinformatics analyses may be performed by a very time and cost-efficient pipeline implemented through VarsVID® on Varstation® online platform.

**Figure 1:**
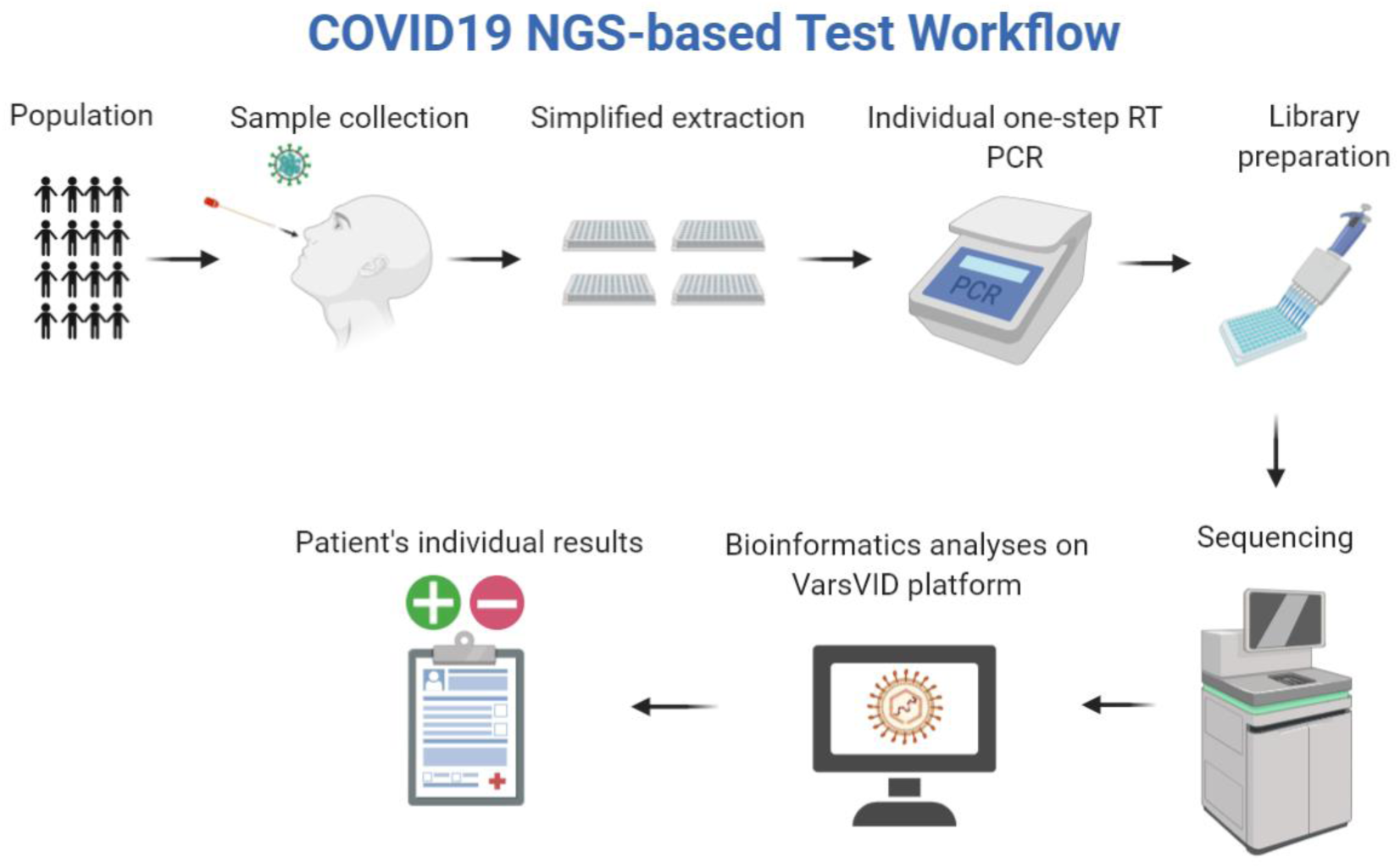
Panoramic view of the COVID19 NGS-based test reported in this work.

### 2.2. Samples and previous results

A total of 269 samples with previous results for COVID19 qRT-PCR were deidentified and selected from routine testing at the Laboratorio de Tecnicas Especiais (LATE), Hospital Israelita Albert Einstein (São Paulo, Brazil). qRT-PCR routine assays were performed using XGEN MASTER COVID-19 kit (Mobius Life Science), which uses a protocol for amplification of fragments of the genes N and ORF1ab. From the 269 samples, 112 tested positive for SARS-CoV-2 with CTs values for gene N ranging from 13 to 35.24. Remaining 157 samples tested negative. We have also included negative plaque-controls with nuclease-free water to assess cross-contamination among plaques or other sequencing artefacts. Finally, AMPLIRUN Coronavirus RNA quantified commercial SARS-CoV-2 control (Vircell Microbiologists, Spain) was serially diluted and tested for assessing limit of detection (LoD). Clinical samples presented in this work were used according to approval of the Hospital Ethical Committee under report #4.178.076.

### 2.3. RNA extractions and internal controls

Total RNA was extracted using the QuickExtract DNA Extraction Solution (Lucigen, WI, USA) according to the manufacturer’s instructions. MS2 phage RNA spike-in was added after extraction to function as an additional routine internal control of true negative samples, i.e., negative samples that do not yield MS2 sequences after bioinformatics analyses will be automatically placed for repetition.

### 2.4. Amplicon generation by RT-PCR

RT-PCR reactions for individual patient samples were performed using SuperScript™ III One-Step RT-PCR (Invitrogen) with a combination of multiplex primers designed to amplify highly conserved regions of SARS-CoV-2 and MS2 control genomes. Several target and control primers were tested for efficiency and multiplex compatibility and sequences are subject to intellectual property. Assays were carried out following standard protocols recommended for the kit: 11 μL template RNA, 2 μL SuperScript™ III RT/Platinum™ Taq Mix (Thermo Fisher Scientific, Waltham, MA), 25 μL 2X Reaction Mix (Thermo Fisher Scientific, Waltham, MA), 1 μL of each primer 10 μM (multiplex primer 1 forward, multiplex primer 1 reverse, multiplex primer 2 forward, multiplex primer 2 reverse), 0.5 μL of internal control primer forward 10 μM, 0.5 μL of internal control primer reverse 10 μM and 7 μL nuclease-free water for a final reaction volume of 50 μL. The RT-PCR reactions were incubated using the following cycling conditions: 56°C for 15 minutes, followed by one cycle of 94°C for 2 minutes, followed by 40 cycles of 94°C for 15 seconds, 65°C for 30 seconds and 72°C for 5 seconds and followed by one cycle of 72°C for 5 minutes.

### 2.5. Library generation

Each patient’s set of individually amplified fragments for SARS-CoV-2 and MS2 genomes was combined in a pool using the Biomek FX Automation (Beckman Coulter). After pool combination, libraries were built using KAPA HiFi Hot Start Enzyme (Roche) and Illumina Nextera XT v2 Index kit set A/B/C/D (Illumina, San Diego, CA, United States). Reaction was carried out with 5 μL cDNA, 25 μL KAPA HiFi Hot Start Enzyme, 5 μL Index i5, 5 μL Index i7, 10 μL nuclease-free water for a final reaction volume of 50 μL. The following cycles were used for the PCR reactions: 95°C for 3 minutes, followed by 8 cycles of 95°C for 30 seconds, 55°C for 30 seconds and 72°C for 5 minutes and followed by one cycle of 72°C for 5 minutes.

### 2.6. Library purification and normalization

Library purification were performed in the Biomek FX Automation (Beckman Coulter) using Agencourt AMPure XP Beads (Beckman Coulter®) following standard protocols. For normalization, all libraries were quantified by Qubit Fluorometer using Qubit™ dsDNA HS Assay Kit (Thermo Fisher Scientific, Waltham, MA) and size assessment was performed on Agilent 2200 TapeStation system for a dilution of 4nM. Each 4nM pool was combined into one pool and submitted to sequencing.

### 2.7. Sequencing

Sample pools were diluted to 2 nM based on the Qiaseq Library Quant Assay Kit (Qiagen) measurements and the size information was performed on Agilent 2200 TapeStation system. For the pool denaturation, 10 μL of the 2 nM pool was added with 10 μL of 0.2 N NaOH, after 10uL de Tris-HCl pH7 200mM was added to stop the denaturation process. 970 μL of Illumina’s HT1 buffer was add to the pool for dilute it to 20 pM and after 117uL of 20 pM pool was add to 1.183uL of Illumina’s HT1 buffer to dilute it to 1.8 pM. Diluted pool was spiked with 15% PhiX and sequenced using a Nextseq 500/550 Mid Reagent Cartridge v2 300 cycles (Illumina, San Diego, CA).

### 2.8. Analyses

Sequencing results were analyzed by a pipeline developed specifically for this highly scalable assay, which performs the general steps of (1) First demultiplexing (2) Quality Control, (3) Mapping to references (4) Assessing reads aligned to targets and (5) Second demultiplexing to generate patient’s individual qualitative results. More specifically, raw sequencing data is demultiplexed using Illumina’s tool blc2fastq with barcode mismatches set to 0 and other parameters set to default. Quality control is performed by cutadapt filtering to size (>50bp), average quality (Q_p_ > 20) and using trimming options to remove NextSeq’s biased low-quality ends. Mapping to references were carried out using bwa-mem tool (default parameters) with SARS-CoV-2 isolate Wuhan-Hu-1 (NC_045512.2) and Phage MS2 (NC_001417.2) as pathogen reference and internal control spike-in, respectively. Reads aligned to target were assessed using Bedtools and qualitative results were generated based on a threshold of reads (≥ 10) and on the decision-table shown in Table 1. A quantitative value was calculated for each positive viral target based on abundance of reads. This metric was termed Fragments per Million (FPM) and is related to similar metrics used in the scientific community for NGS analyses (FPKM, RPKM, etc.). An additional normalization for amplicon length was not necessary, considering that all amplicons were designed to have the same size. FPM values were compared with CT values and Pearson correlation coefficient was calculated. FPM is calculated as follows:

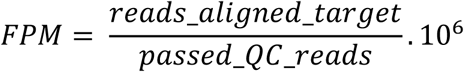

**Table 1:** Decision table based on bioinformatics outputs to generate a patient’s individual qualitative result. Each control or target is considered positive (+) if count of sequences is higher than a selected threshold. Routines samples are placed to repetition when no internal control or target sequences are generated.

| Result | Control | Viral target 1 | Viral target 2 |
| --- | --- | --- | --- |
| Detected | + | + | + |
| Detected | - | + | + |
| Detected | + | + | - |
| Detected | + | - | + |
| Not detected | + | - | - |
| Repeat | - | - | - |

## 3. Results and Discussion

### 3.1. Performance results

In this work, we show a new amplicon-based methodology to amplify highly conserved regions of the SARS-CoV-2 genome and to identify these fragments using NGS sequencing. A total of 269 clinical samples with previous results for COVID19 RT-PCR were tested and performance results are summarized as follows. From 112 samples which were SARS-Cov-2 positive in RT-PCR, 102 were confirmed by NGS. From 157 samples RT-PCR that tested negative, NGS results confirmed 156. These numbers show that the NGS-based technique has overall accuracy of 95.9%, with 91.1% and 99.4% of sensitivity and specificity, respectively. Based on these results we also calculated the positive predictive value (PPV) and the negative predictive value (NPV). PPV value was 99% (102 NGS true positive and 1 NGS false positive) and NPV value was 94% (156 true negatives and 10 false negatives) (Figure 2). We noted that most false negative results presented RT-PCR CT values higher than 30 (6 false negatives). Sensitivity and specificity for samples with CT values lower than 30 were 96.1% and 99.4%, respectively. Accuracy for samples with CT values lower than 30 hitched 98,1% (Figure 2). Average SARS-CoV-2 target NGS coverage for positive samples was 25,172x.

**Figure 2:**
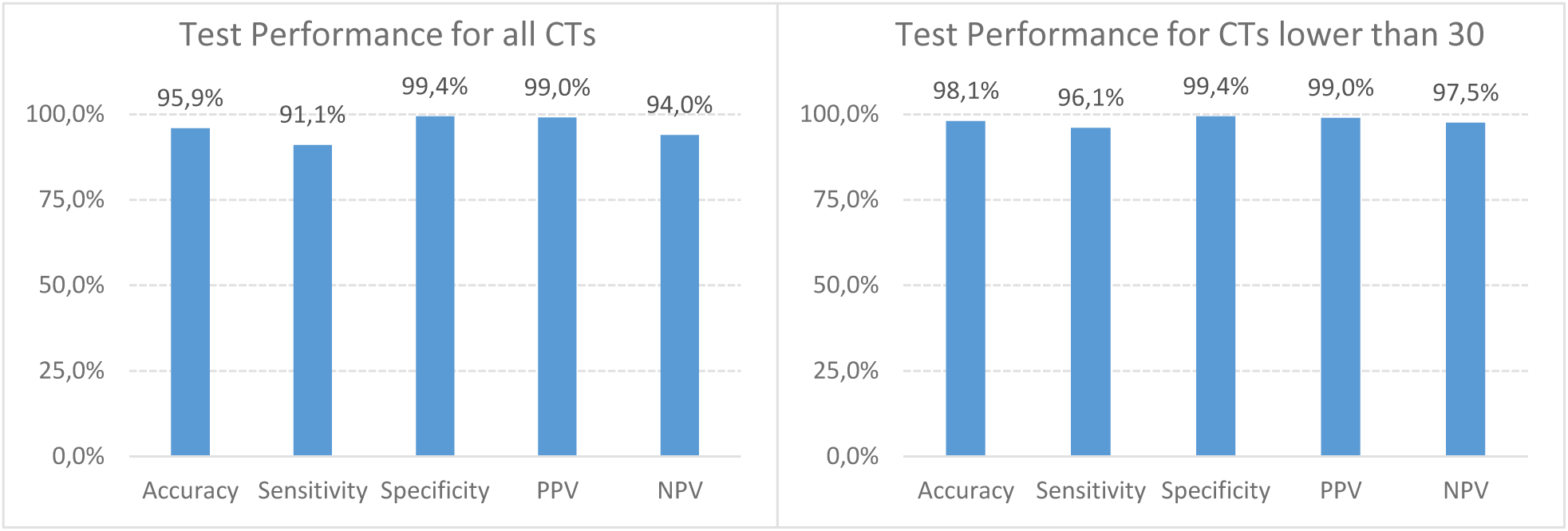
Accuracy, sensitivity, specificity, PPV and NPV percent of SARS-CoV-2 NGS based tested for all samples and for samples with gene N CT values lower than 30.

Positive samples presented average FPM of 2.14 × 10^5^, while average CT for gene N was 24.4. Pearson correlation coefficient between values of CT and FPM was PCC=-0.89 (p-value < 0.001), suggesting that FPM may very well reflect CT values. FPM values of the quantified spike-in controls presented low variance among samples tested, averaging 1.42 × 10^8^ with standard deviation of ± 0.58 × 10^8^. Therefore, we propose this new metric to be associated with qualitative NGS results in the same way as CT values are associated with RT-PCR. FPM values may provide patient and physician an important semi-quantitative idea of viral loads to guide clinical responses. It is important to note, though, that FPM values may vary according to the viral target (Spike, N, Matrix, etc.) and that one target may be more sensitive than others as occurs with RT-PCR.

NGS testing of the AMPLIRUN Coronavirus RNA quantified commercial SARS-CoV-2 control (Vircell Microbiologists, Spain), shows that dilutions from 1 × 10^3^ copies/mL to 1 × 10^5^ copies/mL were successfully identified in replicates by the technique presented in this work. Therefore, we defined 1 × 10^3^ copies/mL as the limit of detection.

### 3.2. Automation and user-friendly results using the VarsVID® platform

As this test was intended to process thousands of samples per run, we anticipated an enormous struggle to organize and perform wet-lab assays. To address this potential bottleneck, we have developed an online platform called VarsVID® to automate the entire process of sample registrations and generation of worksheets. Partner laboratories using VarsVID® will only need to provide a csv file with sample relevant information in order to plan a run and generate worksheets. These worksheets were intended to help the wet-lab professional and to avoid planning mistakes. VarsVID® will also provide a SampleSheet for Illumina sequencing, so that the professional will not waste unnecessary time registering hundreds of libraries in the basespace platform.

Bioinformatics analyses may also be performed using the VarsVID® platform, which was planned to be cloud-based and highly elastic according to laboratory requirements. These efforts resulted in the analyses of hundreds of samples FASTQs in a matter of minutes. Finally, VarsVID® provides user-friendly online results in a dash board, as well as the possibility of generating csv files for further processing.

## 4. Conclusion

While there is a global struggle to control the SARS-CoV-2 pandemic, it is fundamental that new diagnostic methodologies that allows mass testing are made available. Our test was developed considering the best way to guarantee access to accurate molecular diagnosis, without compelling to bottle-necks of supplies such as RNA extraction kits. This proposed methodology uses simple, inexpensive and commonly available reagents for extraction and PCR. Moreover, NGS sequencing costs are extremely diluted when considering the high capacity for sample multiplexing. Finally, VarsVID® platform facilitates the managements of many samples, execution of complex tasks and inspections of results. Altogether, this newly developed NGS-based test is able to detect SARS-CoV-2 with high accuracy, higher scale and lower costs when compared with RT-PCR. We expect that NGS-based testing for COVID19 will multiply world’s capacity of detecting SARS-CoV-2 and fighting this pandemic.

## Data Availability

NGS data, as well as sample sheets and results are available under request.

## Acknowledgements

We thank Dr Cristovao Mangueira and the technical staff of Laboratorio de Tecnicas Especiais, Hospital Israelita Albert Einstein, for scientific and technical support.

## Author contributions

FMM, DA, FCV and MCC conceived and planned this study. BMCA, MSB, COSA, MSN and FMM conducted wet-lab validation. DA and MCC conducted bioinformatics analyses. RAS and JRRP guided and provided insights to the development of this test. All authors have read and contributed to this manuscript.

## References

1. Zhu N, Zhang D, Wang W, Li X, Yang B, Song J, et al. A Novel Coronavirus from Patients with Pneumonia in China, 2019. N Engl J Med. 2020;382(8):727–33.

2. Lu R, Zhao X, Li J, Niu P, Yang B, Wu H, et al. Genomic characterisation and epidemiology of 2019 novel coronavirus: implications for virus origins and receptor binding. Lancet. 2020;395(10224):565–74.

3. Su S, Wong G, Shi W, Liu J, Lai ACK, Zhou J, et al. Epidemiology, Genetic Recombination, and Pathogenesis of Coronaviruses. Trends Microbiol. 2016;24(6):490–502.

4. Coronaviridae Study Group of the International Committee on Taxonomy of V. The species Severe acute respiratory syndrome-related coronavirus: classifying 2019-nCoV and naming it SARS-CoV-2. Nat Microbiol. 2020;5(4):536–44.

5. Sohrabi C, Alsafi Z, O’Neill N, Khan M, Kerwan A, Al-Jabir A, et al. World Health Organization declares global emergency: A review of the 2019 novel coronavirus (COVID-19). Int J Surg. 2020;76:71–6.

6. Wang D, Hu B, Hu C, Zhu F, Liu X, Zhang J, et al. Clinical Characteristics of 138 Hospitalized Patients With 2019 Novel Coronavirus-Infected Pneumonia in Wuhan, China. JAMA. 2020.

7. Lake MA. What we know so far: COVID-19 current clinical knowledge and research. Clin Med (Lond). 2020;20(2): 124–7.

8. Nextstrain. Genomic epidemiology of novel coronavirus - Global subsampling [Available from: https://nextstrain.org/ncov/global.

9. Gohl DM, Garbe J, Grady P, Daniel J, Watson RHB, Auch B, et al. A Rapid, Cost-Effective Tailed Amplicon Method for Sequencing SARS-CoV-2. biorxiv. 2020.

